# Routine Urinary Biochemistry Does Not Accurately Predict Stone Type Nor Recurrence in Kidney Stone Formers: A Multi-Centre, Multi-Model, Externally Validated Machine-Learning Study

**DOI:** 10.1101/2022.06.24.22276866

**Authors:** Robert M Geraghty, Ian Wilson, Eric Olinger, Paul Cook, Susan Troup, David Kennedy, Alistair Rogers, Bhaskar K Somani, Nasser A Dhayat, Daniel G Fuster, John A Sayer

**Affiliations:** Department of Urology, Freeman Hospital, Newcastle-upon-Tyne; Translational and Clinical Institute, Faculty of Medical Sciences, Newcastle University, Central Parkway, Newcastle upon Tyne, England, United Kingdom; Biosciences Institute, Newcastle University, International Centre for Life, Central Parkway, Newcastle-upon-Tyne; Department of Biochemistry, University Hospital Southampton, Southampton; Department of Biochemistry, Queen Elizabeth Hospital, Gateshead; Department of Urology, University Hospital Southampton, Southampton; Department of Nephrology and Hypertension, Inselspital, Bern University Hospital, University of Bern, Switzerland; Department of Nephrology and Hypertension, Inselspital, Bern University Hospital, Department for Biomedical Research, University of Bern, Switzerland; The Newcastle upon Tyne Hospitals NHS Foundation Trust, Freeman Road, Newcastle upon Tyne, England, United Kingdom; National Institute for Health Research Newcastle Biomedical Research Centre, Newcastle upon Tyne, England, United Kingdom

**Keywords:** Kidney stones, Machine learning, Prediction, Urinary Biochemistry

## Abstract

**Objectives:** Urinary biochemistry is used to detect and monitor conditions associated with recurrent kidney stones. There are no predictive machine learning (ML) tools for kidney stone type or recurrence. We therefore aimed to build and validate ML models for these outcomes using age, gender, 24-hour urine biochemistry and stone composition.

**Materials and Methods:** Data from 3 cohorts were used, Southampton, UK (n=3013), Newcastle, UK (n=5984) and Bern, Switzerland (n=794). Of these 3130 had available 24-hour urine biochemistry measurements (calcium, oxalate, urate, pH, volume), and 1684 had clinical data on kidney stone recurrence. Two predictive models were constructed (UK and Swiss) using two ML techniques (Partitioning and Random Forests [RF]) and validated internally with a subset of the same dataset (e.g UK model/UK test set), and externally with the other dataset (UK model/Swiss test set).

**Results and Limitations:** For kidney stone type, on external validation accuracy of UK RF model=0.79 (95% CI: 0.73-0.84), sensitivity: calcium oxalate=0.99 and calcium phosphate/urate=0.00. Specificity: calcium oxalate=0.00 and calcium phosphate/urate=0.99. For the Swiss RF model accuracy=0.87 (95% CI: 0.83-0.89), sensitivity: calcium oxalate=0.99 and calcium phosphate/urate=0.00. Specificity: calcium oxalate=0.00, calcium phosphate=0.00 and urate=1.00.

For stone recurrence, on external validation accuracy of UK RF model=0.22 (95% CI: 0.19-0.25), sensitivity=0.93 and specificity=0.09. Swiss RF model accuracy=0.42 (95% CI: 0.39-0.47), sensitivity=0.03 and specificity=0.97.

**Conclusions:** Neither kidney stone type nor kidney stone recurrence can be accurately predicted using modelling tools built using specific 24-hour urinary biochemistry values alone. Further studies to delineate accurate predictive tools should be undertaken using both known and novel risk factors.

## Introduction

Kidney stones are prevalent^1^, costly^2^ and debilitating, with a recurrence rate of around 40-50% at 10 years^3^. Although there are rare well described forms of kidney stones such as cystinuria leading to cystine stones, the pathophysiology underlying the most common types of kidney stones (calcium stones and urate) have not been satisfactorily described. Given the rising prevalence and cost of surgical treatment, tools are needed to accurately predict kidney stone type and more importantly, risk of recurrence so that preventative measures can be more focused and beneficial.

Kidney stone composition may not always be investigated, but remains vital for the identification of rarer stone types that require further management. There has previously been only one machine learning (ML) study that looked at prediction of kidney stone type^4^. Several types of ML models were constructed for kidney stone type prediction, achieving high accuracy (area under the curve (AUC)=0.99), although it was neither internally nor externally validated. They did not use urinary biochemistry, instead utilising demographics and basic medical history. Another study applied logistic regression to 24-hour urinary biochemistries to predict stone type, but this demonstrated poor predictive value^5^. Regression models are not as powerful as more modern ML techniques^6^.

The Recurrence of Kidney Stone (ROKS) nomogram was created to predict kidney stone recurrence and is largely based on non-biochemical risk factors and radiologic appearances, rather than biochemistry^7^. It had a moderate predictive value (AUC=0.65) for kidney stone recurrence at 2-years on external validation^8^. Until accuracy improves, this predictive model is not useful for influencing clinical practice.

Twenty four hour urine samples are often taken from high-risk stone formers^9^ to identify stone forming risk factors such as hyperoxaluria, hypercalciuria, hyperuricosuria etc. Although the benefit of urinary biochemistry for managing kidney stone patients has not been fully established^10^, there is evidence from randomized controlled trials that medications used in specific, limited circumstances reduce the number and increase the time to, recurrence^11^. A systematic review by Hsi et al. delineates the limitations of 24-hour urine collection, none of the included studies demonstrate any predictive value for recurrence^10^.

To date there have been no predictive ML models for kidney stone type or recurrence based on 24-hour urinary biochemistry. We therefore aimed to construct predictive ML models for these outcomes using clinical and biochemical data from kidney stone patients to assess the predictive value of 24-hour urinary biochemistry.

## Methods

### Data Collection across all Cohorts

Age, sex, kidney stone recurrence, stone type and 24-hour urinary biochemistry (free-choice diet).

Urinary biochemistry obtained: calcium (mmol/24 hour), oxalate (mmol/24 hour), urate (mmol/24 hour), urine volume (L) and pH. Citrate (mmol/24 hour) was obtained from the Newcastle and Bern cohorts.

Stone type was ascertained using Fourier Transform IR spectroscopy in all cohorts. Differentiation between calcium oxalate types (monohydrate/dihydrate) was not available in the Southampton/Newcastle cohorts and therefore not included in analysis.

### Southampton, UK

#### Study Population

The cohort consisted of patients with kidney stone disease presenting to University Hospital Southampton, UK (tertiary referral hospital) referred for metabolic assessment between 1990-2007. The initial cross-sectional study (n=2801)^12^ and subsequent cohort subset^2^ have been described. During this period, stone formers were routinely referred from across Hampshire, Isle of Wight and Dorset. The subset includes 1000 patients with further information ascertained retrospectively using hospital/general practice electronic records.

Patients who had no documentation (i.e no evidence of subsequent follow-up/consultation, lived outside/have left Hampshire or no documentation on CHIE) were excluded [see fig. 1].

**Figure 1.**
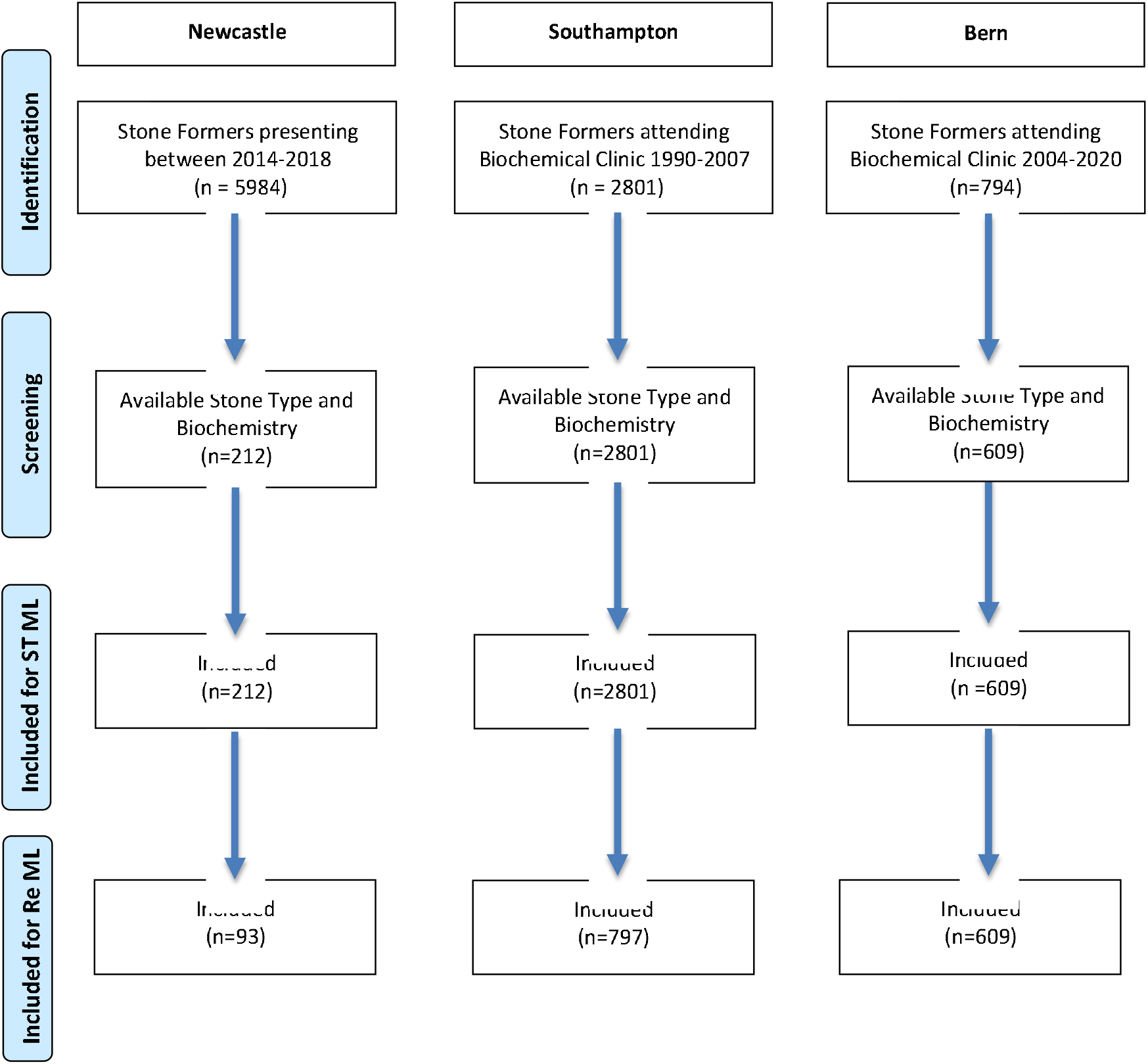
Flow diagram of patient selection/Inclusion. ML=Machine Learning, ST=Stone type, Re=Recurrence.

#### Ethical Approval

Ethical approval for this arm of the study was granted by the NHS Bristol Research Ethics Committee (Rec ref: 18/SW/0185; IRAS ID: 240061).

### Newcastle upon Tyne, UK

#### Study Population

The cohort consisted of patients with kidney stone disease presenting to hospitals in the north of England between 2013 and 2018 [Appendix 1]. These include: The Newcastle upon Tyne Hospitals NHS Foundation Trust (Newcastle-upon-Tyne), James Cook University Hospital (Middlesbrough), Sunderland Royal Hospital, Durham University Hospital, Queen Elizabeth Hospital (Gateshead), Darlington Memorial Hospital, Cumberland Infirmary (Carlisle) and Northumbria Hospitals Trust (Wansbeck, Hexham, Cramlington).

Data ascertained retrospectively using electronic hospital records. Urinary biochemistry and stone type were obtained from the regional renal calculi processing unit (Queen Elizabeth Hospital, Gateshead).

#### Ethical Approval

This arm was registered as an audit (current patients of authors) with Newcastle-upon-Tyne Hospitals NHS Foundation Trust (audit number: 10337).

### Bern, Switzerland

#### Study Population

Patients enrolled in the Bern Kidney Stone Registry (BKSR) were referred to the Department of Nephrology and Hypertension, Inselspital, Bern University Hospital and included kidney stone formers suffering at least one stone episode. This population has previously been described in the literature^13^. Those <18 years were excluded.

#### Ethical Approval

The BKSR adheres to the Declaration of Helsinki, was approved by the Ethical Committee of the Kanton Bern (#95/06) and all patients provided written informed consent.

### Definitions

Stone recurrence was defined as subsequent stone surgery or acute presentation following previous treatment/spontaneous passage. Presence of asymptomatic recurrence on imaging was not available.

Low urine volume was defined as <1.5L/24 hour. Acid urine was defined as pH <5.5.

Biochemical abnormalities were defined as follows, as per local laboratory values: hypercalciuria in women >6.2 mmol/24 hour and men >7.5 mmol/24 hour; hyperoxaluria in women >0.32 mmol/24 hour, men >0.49 mmol/24 hour; hyperuricosuria as >4.46 mmol/24 hour and hypocitraturia as <0.6 mmol/24 hour.

Primary kidney stone type was defined as >50% of particular type.

### Statistical Analysis

Analysis was performed in R (version no.4.0.2). For robustness, two machine learning models were used: Partitioning (rpart[19]) and Random Forests (randomForest[20] and missForest[21]) for both Stone Type and Recurrence. Missing data was accounted for with values imputed from existing data (missForest). Variables included in both analyses: age, sex, raw 24-hour urinary values (calcium, oxalate, urate, volume), pH and number of biochemical abnormalities. Urinary citrate was excluded due to low numbers with documented citrate. To investigate whether unbalanced data was driving discrimination, up-sampling of training data was used to create balanced training data sets using caret[22], which were run through subsequent analyses.

Recurrence rate was excluded in stone type models as this would be unknown at point of presentation. Stone type was included in the recurrence models. Those with unclear stone type were excluded from the Stone Type models. Data was partitioned into training (70% of total) and test (30%) datasets. Diagnostic accuracy statistics are presented in the results section. Sample sizes were not calculated as an effect size was not being calculated. For robustness, two models were used for each measure, and models were constructed from two cohorts. Sensitivity analysis is represented by the second model (random forests), and by the second dataset (Bern).

## Results

### Patient and Biochemistry Demographics

There were 3013 kidney stone patients in the Southampton/Newcastle combined dataset [see table 1]. There were 2074 men (mean age 49±15 years) and 939 women (mean age 49±17 years). Stone types were as follows: calcium oxalate (n=1476, 49%), calcium phosphate (n=119, 4%), urate (n=89, n=3%), cystine (n=5, 0.2%), struvite (magnesium ammonium phosphate) (n=3, 0.1%) and no data (n=1321, 44%). Those with no data are likely to have passed their stone/post treatment fragments (e.g. shockwave lithotripsy) spontaneously. The stone was then not available for analysis. Of these 3013, 890 had information on whether or not the patient had a kidney stone recurrence. Of these, 374 had a recurrence (39%). The low numbers of patients with follow-up information are likely due to the patient not accessing medical services following the initial stone episode, moving away or dying.

**Table 1.** Patient and Kidney Stone Demographics. Age in years.

|  |  | Southampton | Newcastle | Combined<br>Southampton &<br>Newcastle | Bern |
| --- | --- | --- | --- | --- | --- |
| **Total, n** |  | 2801 | 212 | 3013 | 794 |
| **Sex** | **Female, n** | 818 (29%) | 91 (43%) | 939 (31%) | 204 (26%) |
|  | **Female, Age±SD** | 49±16 | 50±19 | 49±17 | 45±15 |
|  | **Male, n** | 1984 (71%) | 121 (57%) | 2073 (69%) | 590 (74%) |
|  | **Male, Age±SD** | 49±14 | 48±16 | 49±15 | 48±13 |
| **Stone Type** | **Calcium Oxalate, n (%)** | 1393 (50%) | 83 (39%) | 1476 (49%) | 598 (75.9%) |
|  | **Calcium Phosphate, n (%)** | 59 (2%) | 60 (28%) | 119 (4.0%) | 81 (10.3%) |
|  | **Urate, n (%)** | 55 (2%) | 34 (16%) | 89 (3.0%) | 47 (6.0%) |
|  | **Cystine, n (%)** | 5 (0.2%) | 0 | 5 (0.2%) | 16 (2.0%) |
|  | **Struvite, n (%)** | 1 (0.04%) | 2 (1%) | 3 (0.1%) | 6 (0.8%) |
|  | **No data, n (%)** | 1288 (46%) | 33 (16%) | 1321 (44%) | 40 (5.1%) |
| **Recurrence, n/total with data (%)** |  | 325/855 (38%) | 49/111 (44%) | 374/890 (39%) | 676/794 (85.1%) |

There were 794 kidney stone patients in the Bern dataset, with 204 women (mean age 45±15 years) and 590 men (mean age 48±13 years). Stone types were as follows: calcium oxalate (n=598, 76%), calcium phosphate (n=81, 10%), urate (n=47, n=6%), cystine (n=16, 2%), struvite (n=6, 0.8%) and no data (n=40, 5.1%) [see fig 2]. All had follow-up data available for stone recurrence. There were 676 recurrences (85%).

**Figure 2.**
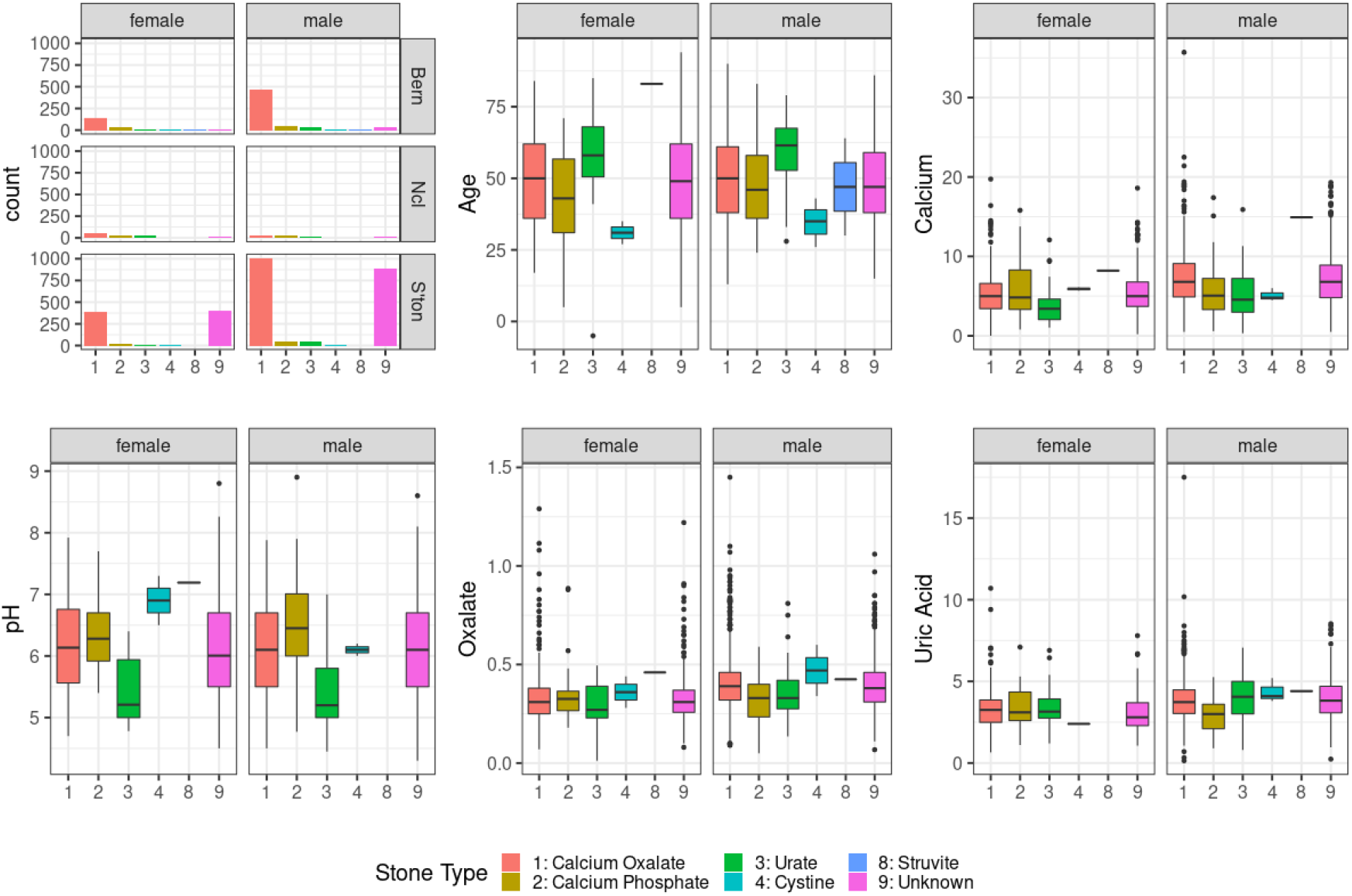
Graphical representation of differences between cohorts and sexes. On top row, from left to right: bar chart of number of patients in each sex, in each cohort by stone type; boxplot of age for each sex per stone type; boxplot of 24-hour urinary calcium (mmol/24 hour) for each sex per stone type. Bottom row, from left to right: boxplot of urinary pH for each sex per stone type; boxplot of 24-hour urinary oxalate (mmol/24 hour) for each sex per stone type; boxplot of 24 hour urinary uric acid (mmol/24 hour) for each sex per stone type.

Across all 3 datasets, 3130 patients had full data on urinary biochemistry. There were 631 (20.1%) patients who had no biochemical diagnoses, 1331 (42.5%) patients with a single diagnosis and 1668 (37.3%) with a combination of biochemical abnormalities.

Of those with full data on urinary biochemistry, 1002 patients had data on kidney stone recurrence. In total 688 (68.7%) had a recurrence. There were 272 (74.7%) recurrences in those with no biochemical diagnosis, 258 (67.0%) with a single biochemical risk factor and 163 (64.4%) in those with a combination [see appendices 2 & 3].

The Southampton dataset did not contain 24-hour urinary citrate measurements. In the Newcastle & Bern datasets there were 45 and 121 patients, respectively, with hypocitraturia. Recurrence data was available in 144 patients, with recurrence in 11/23 (Newcastle, 47.8%) and 109/121 (Bern, 90.0%), respectively.

Discordant primary kidney stone type and 24-hour urine biochemistry was seen in 103 patients (5% of known stone type). Cystine stones (n=21) and struvite stones (n=9) were not included in the appendices given their low numbers and alternative aetiologies.

Data on subsequent primary stone type was available in the Bern dataset for a subset of those with a stone recurrence (n=188). Change in primary stone type from initial episode was seen in 42 patients (22%): calcium oxalate (n=26, 18%), calcium phosphate (n=10, 37%), urate (n=5, 38%), cystine (n=0, 0%), struvite (n=1, 100%) [Figure 3].

**Figure 3.**
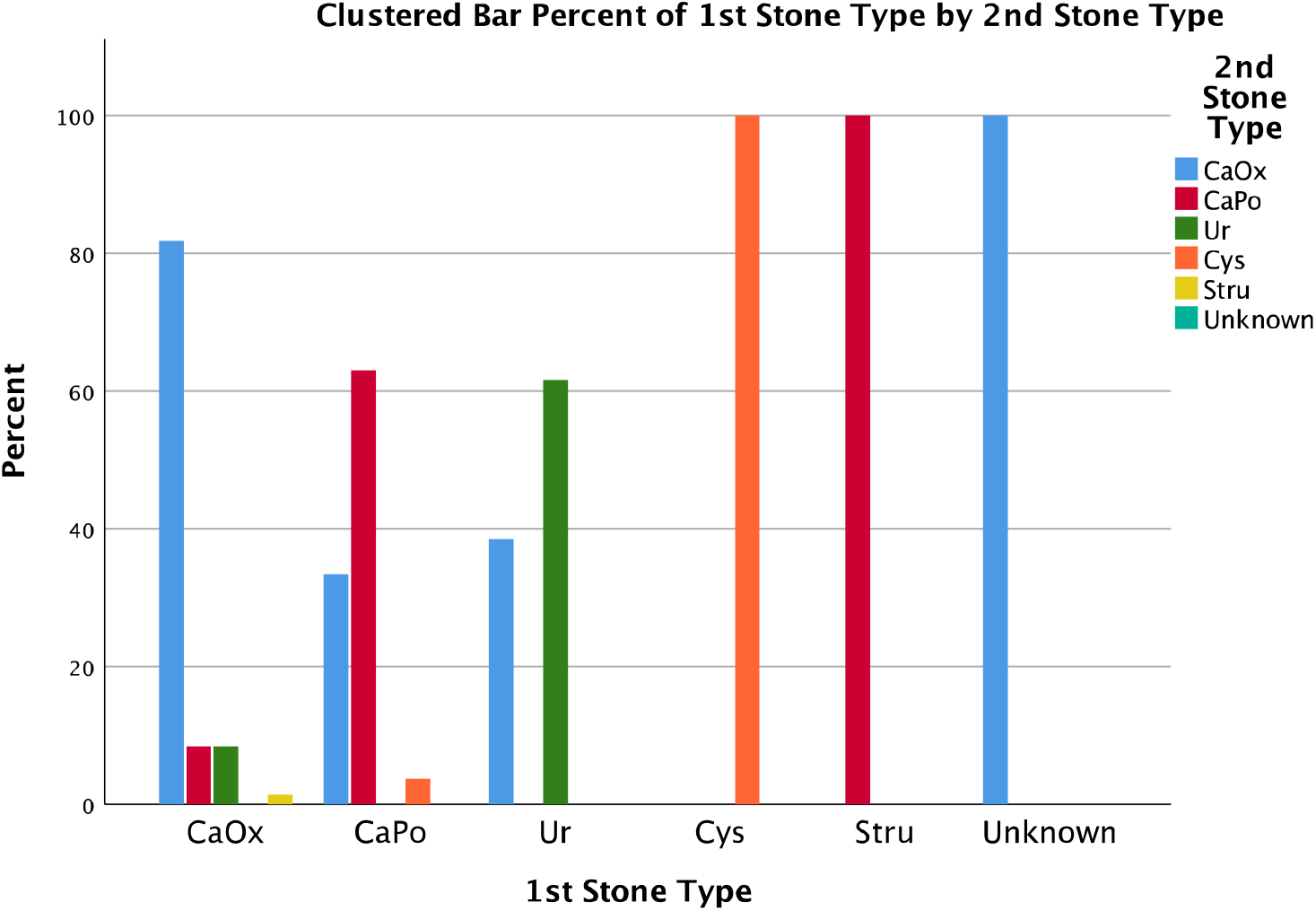
Bar chart showing percentage of subsequent stone type for each first stone type, data from the Bern Cohort. CaOx=Calcium Oxalate, CaPo=Calcium Phosphate, Ur=Urate, Cys=Cystine, Stru=Struvite

### Machine Learning Models

#### Stone Type

The training group had n=3092 patients, n=2360 of which had immediate clearance on fluoroscopy. On internal validation (test set: n with outcome/total; n=1015/1326), the diagnostic accuracy statistics were: Random forests (RF) AUC=0.73, accuracy= 0.77 (95% CI:0.74-0.79), sensitivity=0.27, specificity=0.94; Partitioning AUC=0.63, accuracy=0.71(95% CI:0.69-0.74), sensitivity=0.22, specificity=0.89; Extreme Gradient Boosting (XGBoost) AUC=0.75, accuracy=0.77 (95% CI:0.75-0.80), sensitivity=0.27, specificity=0.94; Logistic Regression (LR) AUC=0.63, accuracy=0.76 (95% CI:0.73-0.78), sensitivity=0.12, specificity=0.97; Classical Neural Network (NN) AUC=0.74, accuracy=0.75 (95% CI:0.73-0.78), sensitivity=0.23, specificity=0.94; Bayesian GeneralisedLinear Model (BGLM) AUC=0.75, accuracy=0.77 (95% CI:0.75-0.80), sensitivity=0.29, specificity=0.94; Deep Neural Network (DNN) Single-outcome model AUC=0.59, accuracy=0.77 (95% CI:0.75-0.79), sensitivity=0.57, specificity=0.79; DNN Multiple-outcome model AUC=0.53, accuracy=0.77 (95% CI:0.75-0.79), sensitivity=0.60, specificity=0.78 [see figure 2 and supplementary table 2].

**Table 2.** Descriptive statistics for Partitioning and Random Forests models predicting recurrence. PPV=positive predictive value, NPV=negative predictive value. Test set=30% of same cohort

| Recurrence | Southampton/Newcastle Model |  | Bern Model |  |
| --- | --- | --- | --- | --- |
|  | Partitioning | Random Forests | Partitioning | Random Forests |
| **Accuracy (95% CI)** | 0.52 (0.45-0.60) | 0.55 (0.48-0.63) | 0.85 (0.80-0.89) | 0.89 (0.84-0.93) |
| **Sensitivity** | 0.64 | 0.69 | 0.07 | 0.05 |
| **Specificity** | 0.35 | 0.35 | 0.95 | 0.99 |
| **PPV** | 0.60 | 0.61 | 0.15 | 0.33 |
| **NPV** | 0.40 | 0.42 | 0.89 | 0.90 |
| **Detection Rate** | 0.39 | 0.41 | 0.008 | 0.005 |

Two models were built to predict kidney stone type, partitioning and Random Forests (RF). Both models used all available 24 hour urinary biochemical data (volume, calcium, oxalate, urate and pH), age and sex. Citrate was excluded as the Southampton data did not have this. Due to missing values (as detailed above), some biochemical values were imputed. Up-sampling of rare stone data, while marginally improving classification into rare stone types, overall gave inferior classification, therefore simple training sets are reported.

For the internally validated Southampton/Newcastle models, the partitioning model accuracy was 0.89 (95% CI: 0.85-0.91) and RF model accuracy was 0.90 (95% CI: 0.87-0.92) [see table 3].

For the internally validated Bern models, the partitioning model accuracy was 0.75 (95% CI: 0.69-0.81) and RF model was 0.81 (95% CI: 0.78-0.84).

None of the models were able to predict cystine/struvite stones.

Each RF model was externally validated using the other dataset. For the Southampton/Newcastle model, validated with Bern data, the accuracy was 0.79 (95% CI: 0.73-0.84). The sensitivity for: calcium oxalate=0.99, calcium phosphate/urate=0.00. The specificity for: calcium oxalate=0.00 and calcium phosphate/urate=0.99.

For the Bern model, validated with Southampton/Newcastle data, the accuracy was 0.87 (95% CI: 0.83-0.89). The sensitivity for: calcium oxalate=0.99, calcium phosphate=0.00 and urate=0.04. The specificity for: calcium oxalate=0.02, calcium phosphate=0.99 and urate=1.00.

Addition of citrate into the models with the Newcastle and Bern datasets did not demonstrate any difference in accuracy, sensitivity or specificity for stone type. Increasing the predominant stone type threshold for calcium phosphate stones to 90% did not increase accuracy, sensitivity or specificity.

#### Recurrence

Two models were used to predict recurrence (Partitioning and RF). Both models used all biochemistry previously described, age and sex. Biochemical value imputation as above.

For the internally validated Southampton/Newcastle models, the partitioning model accuracy was 0.52 (95% CI: 0.45-0.60), correctly predicting 25/72 recurrences and 70/109 without. The RF model accuracy was 0.55 (95% CI: 0.48-0.63), correctly predicting 25/72 recurrences and 75/109 without [see Table 4].

For the internally validated Bern models, the partitioning model accuracy was 0.85 (95% CI: 0.80-0.89), correctly predicting 201/212 recurrences, and 2/27 without. The RF model accuracy was 0.89 (95% CI: 0.84-0.93), correctly predicting 163/165 recurrences, and 1/19 without.

Each RF model was externally validated as above. The Southampton/Newcastle model, validated with Bern data, accuracy was 0.22 (95% CI: 0.19-0.25), sensitivity=0.93, specificity=0.09, correctly predicting 55/632 recurrences and 108/116 without.

The Bern model, validated with Southampton/Newcastle data, accuracy was 0.42 (95% CI: 0.39-0.47), sensitivity=0.03, specificity=0.97, correctly predicting 235/242 recurrences and 11/332 without.

Addition of citrate into the models with the Newcastle and Bern datasets did not demonstrate any difference in accuracy, sensitivity or specificity for stone type. Increasing the predominant stone type threshold for calcium phosphate stones to 90% did not increase accuracy, sensitivity or specificity.

## Discussion

This study utilised 24-hour urinary biochemistry data from kidney stone patients attending three European tertiary referral centres to make two cohorts (Southampton/Newcastle and Bern), used to construct internally validated machine learning (ML) models for both kidney stone type and kidney stone recurrence. These models were then externally validated on the other cohort. The recurrence models had very poor accuracy, whilst the kidney stone type models demonstrated high overall accuracy. However this is likely due to the high number of calcium oxalate kidney stones in our datasets (reflected in the literature [23]), the diagnostic statistics demonstrate very poor predictive value for other stone types. In essence the models predict every stone as calcium oxalate.

The poor predictive value for both outcomes are important negative findings. The composition of most kidney stones is unknown (36% in this study and ≤85% in the literature[24]), and 24-hour urine collection is used to identify recurrent/high-risk stone formers who could be started on prophylaxis [11,12].

The main strengths of this study are the large cohort sizes, internal & external validation[19,20], multiple ML models (superior to regression analyses[8]) showing similar outcomes and consistent outcomes across both datasets.

There are several weaknesses with this study. Few of the Newcastle kidney stone patient cohort had 24-hour urinary biochemical analysis available, and fewer (in both datasets) had data on recurrence. This was addressed with imputation of missing data[21]. Unfortunately, 24-hour urinary citrate was not available in the Southampton dataset, which may bias our main results as hypocitraturia is an important risk factor for calcium-containing kidney stones[25]. To investigate this both models were repeated with citrate included for the Bern/Newcastle datasets, this did not demonstrate any improvement in diagnostic statistics. The datasets did not include 24-hour urinary phosphate or sodium, both of which influence the risk of stone formation[26]. Therefore, predictive value may be improved with the addition of these variables. The stone type models could not predict struvite or cystine stones. This is likely due to minimal numbers of these stones, thus underpowering the model, and their alternative pathophysiology[27,28]. Although it is accepted that those with cystinuria, are at high risk of recurrence[28].

The datasets differ in terms of stone types proportions, notably uric acid and calcium phosphate stones. For instance, in the Southampton and Bern cohorts uric acid stones represent 3.6% and 6% respectively, whilst in the Newcastle dataset 19% have these (10% of total 5984 patients). Uric acid stones are associated with obesity/metabolic syndrome[29], and this observation may represent rising levels in Newcastle compared to an historic population (Southampton), or a contemporary European population (Bern). This could also represent referral bias, e.g. in the Bern cohort referral rates for urate stones were historically lower than present. As information on BMI or diabetes mellitus was not available this limits the predictive value of the models to predict stone type.

Proportions of calcium phosphate stones are also different between datasets (2% in Southampton, whilst 28% in Newcastle). This may be due to selection bias, as large numbers of patients in the Newcastle set did not have urinary biochemistry available. This may also be due to our definition of predominant stone type (i.e >50%). Pure calcium phosphate stones (>90%) are associated with a specific phenotype (alkaline urine and hypocitraturia) [30]. Increasing the cut-off for calcium phosphate stones to >90% did not demonstrate an increase in predictive power for either stone type or recurrence. This may be due to the low numbers with this stone type (UK: from 4% to 2.8%, Bern: 10.3% to 2.6%). This touches on the wider issue of stone type interpretation. There are no guidelines on how to interpret stone composition, nor are there consistent methods within the literature. It is well known that stones can take many forms/compositions, but translation of these observations has yet to reach everyday practice. Our use of a >50% cut-off was an attempt to use standard laboratory outputs to improve model accessibility.

As well as differences in stone type, recurrence data also differed between datasets, Bern had a far higher rate than the UK cohorts (which are similar to the literature[5]). This is likely due to the selection of patients going to biochemical clinic/tertiary care and will likely skew the results as these patients are not representative of the general stone forming population. This is reflected in the recurrence models, the Bern model predict most as recurrence, whilst the Southampton/Newcastle model predict most as no recurrence.

24-hour urinary biochemistry is touted as a good measure to identify urinary risk factors for stone formation[11,12]. In certain circumstances this is true[15], however there are a significant minority of recurrent kidney stone formers who have no biochemical abnormalities (20% in these cohorts and similar to rates seen in other studies[14,31]). This may be due to dietary influence on urinary biochemistry, with a random diet unpredictably influencing the results[32]. Further, a small proportion of patients have discordant urinary biochemistry and stone type (3.3% across these cohorts), which makes prediction of unknown stone type unreliable. In summary, the main challenge to model accuracy was not unbalanced data but a lack of factors that could provide effective discrimination.

There is a growing body of evidence that kidney stone disease is the common end point of a heterogenous group of disorders that include environmental[33], dietary[34,35], monogenic[36] and polygenic risk factors[37,38], whether in isolation or combination. Therefore reliance on urinary biochemistry alone as a predictive tool, as this study demonstrates, is inadvisable. This study, along with a recent study demonstrating the futility of serum biochemistry (except calcium)[39], demonstrate that current practices[11,12] to delineate risk of kidney stone recurrence do not work well. At least 15% of kidney stones have a monogenic cause[36], with a significant proportion forming calcium stones[40]. It is evident that a precision medicine approach is needed to differentiate the differing causes of KSD, which may then be treated effectively[41].

Future studies are needed to delineate an effective kidney stone screen based on traditional risk factors in combination with kidney stone analysis, 24-hour urinary biochemistry, monogenic screen and polygenic risk scoring.

## Conclusion

Neither kidney stone type nor kidney stone recurrence can be accurately predicted by 24-hour urinary biochemistry. Further studies to delineate an effective stone screen should be undertaken.

## Data Availability

All data produced in the present study are available upon reasonable request to the authors

## Acknowledgements

Robert Geraghty is a National Institute for Health Research Academic Clinical Fellow. Eric Olinger is supported by an Early Postdoc Mobility Stipendium of the Swiss National Science Foundation and Kidney Research UK. John Sayer is funded by Kidney Research UK and the Northern Counties Kidney Research Fund.

We would like to acknowledge Dr Valerie Walker for initiating the Southampton stone database.

---

**Appendix 1.**
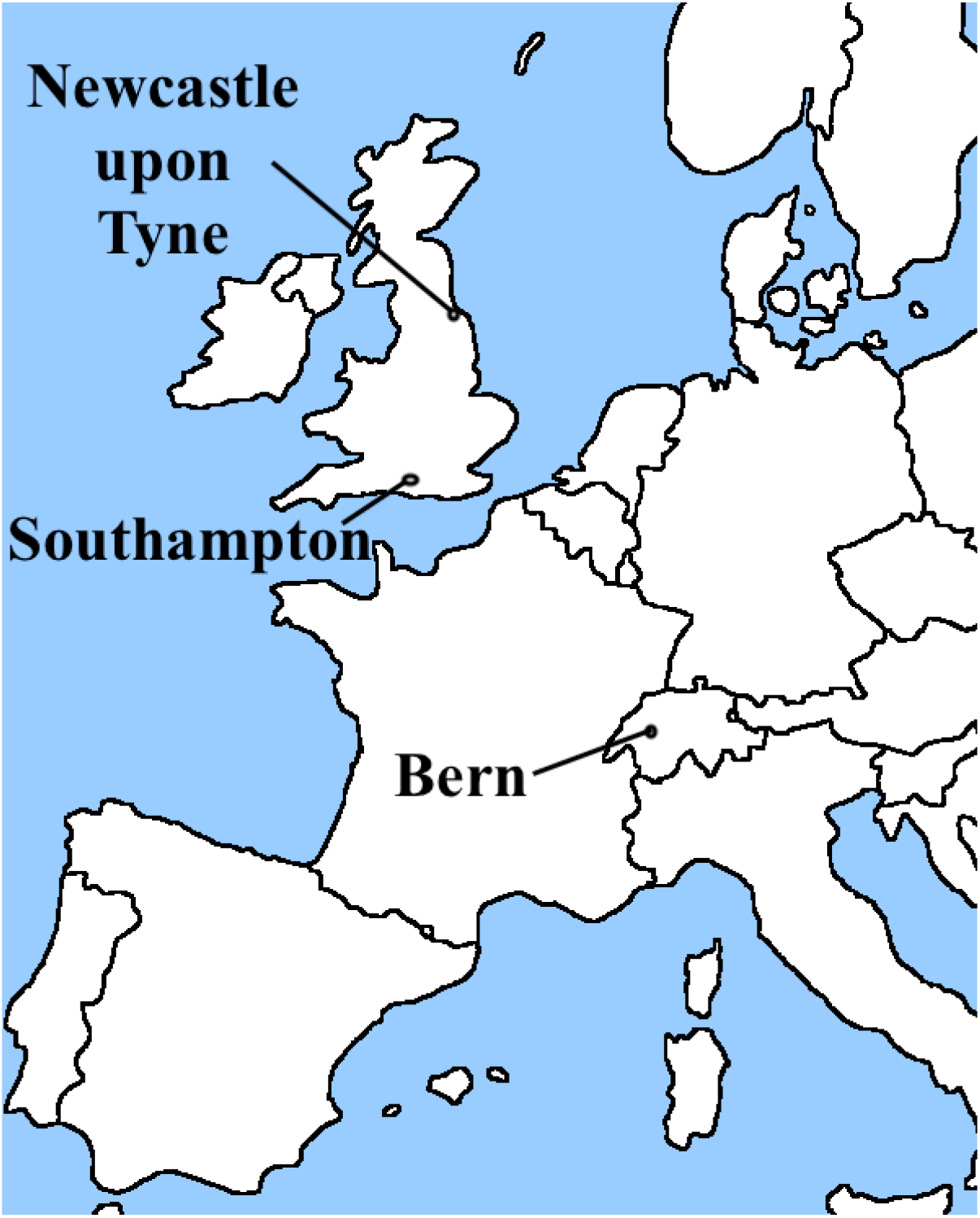
Study centre locations.

**Appendix 2.**
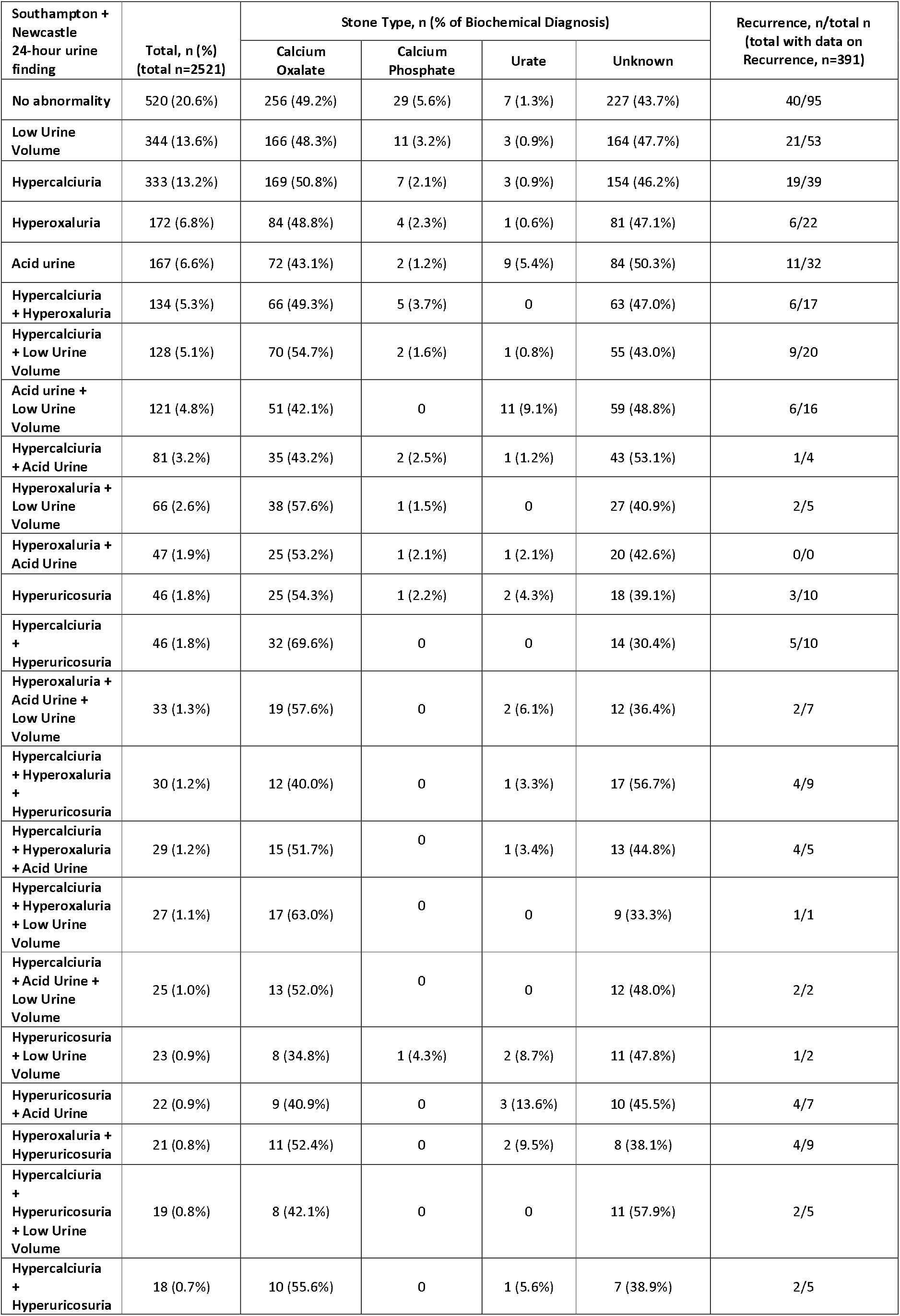
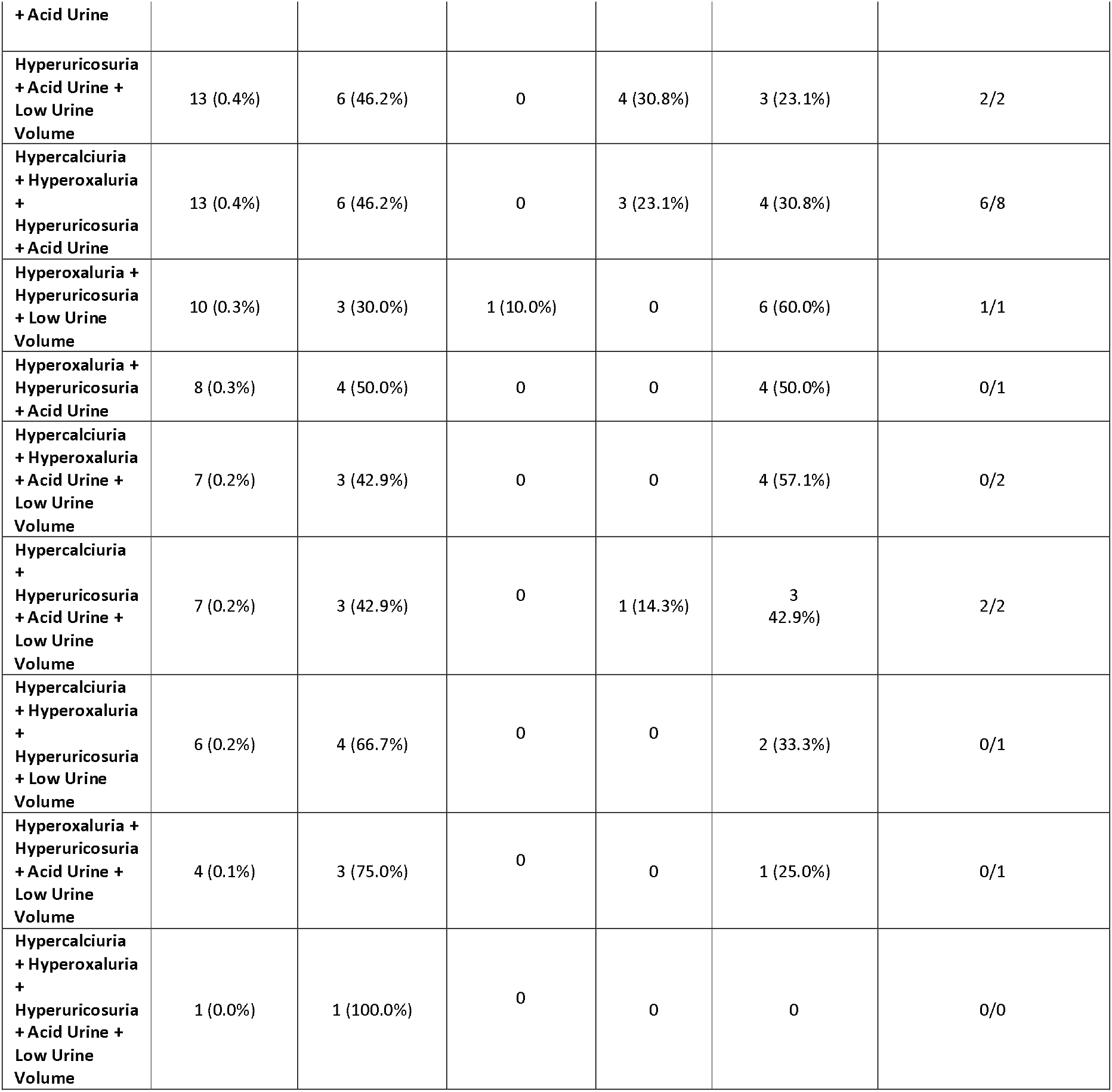
Biochemistry diagnoses and recurrences for Newcastle/Southampton dataset.

**Appendix 3.** Biochemistry diagnoses and recurrences for Bern database.

*Appendix 2. Biochemistry diagnoses and recurrences for Newcastle/Southampton dataset.*
| Bern<br>24-hour urine finding | Total, n<br>(%)<br>(total<br>n=609) | Stone Type, n (% of Biochemical Diagnosis) |  |  |  | Recurrence,<br>n/total n (total<br>with data on<br>Recurrence,<br>n=609) |
| --- | --- | --- | --- | --- | --- | --- |
|  |  | Calcium Oxalate | Calcium<br>Phosphate | Urate | Unknown |  |
| No abnormality | 111<br>(18.2%) | 85 | 13 | 3 | 3 | 97/111 |
| Low Urine Volume | 80<br>(13.1%) | 65 | 5 | 3 | 4 | 66/80 |
| Hypercalciuria | 53<br>(8.7%) | 43 | 7 | 0 | 2 | 46/53 |
| Hyperoxaluria | 78<br>(12.8%) | 50 | 13 | 6 | 4 | 71/78 |
| Acid urine | 50<br>(8.2%) | 36 | 2 | 6 | 4 | 41/50 |
| Hypercalciuria +<br>Hyperoxaluria | 24<br>(3.9%) | 19 | 4 | 0 | 1 | 23/24 |
| Hypercalciuria + Low<br>Urine Volume | 12<br>(2.0%) | 11 | 0 | 0 | 1 | 11/12 |
| Acid urine + Low Urine<br>Volume | 31<br>(7.1%) | 24 | 0 | 5 | 2 | 26/31 |
| Hypercalciuria + Acid<br>Urine | 5 (0.8%) | 5 | 0 | 0 | 0 | 4/5 |
| Hyperoxaluria + Low Urine<br>Volume | 15<br>(2.5%) | 12 | 0 | 1 | 2 | 11/15 |
| Hyperoxaluria + Acid<br>Urine | 32<br>(5.3%) | 26 | 3 | 1 | 2 | 25/32 |
| Hyperuricosuria | 8 (1.3%) | 6 | 1 | 1 | 0 | 8/8 |
| Hypercalciuria +<br>Hyperuricosuria | 15<br>(2.5%) | 12 | 2 | 0 | 1 | 13/15 |
| Hyperoxaluria + Acid<br>Urine + Low Urine Volume | 11<br>(1.8%) | 8 | 0 | 3 | 0 | 11/11 |
| Hypercalciuria +<br>Hyperoxaluria +<br>Hyperuricosuria | 15<br>(2.5%) | 11 | 2 | 0 | 2 | 14/15 |
| Hypercalciuria +<br>Hyperoxaluria + Acid<br>Urine | 9 (1.5%) | 7 | 0 | 1 | 1 | 8/9 |
| Hypercalciuria +<br>Hyperoxaluria + Low Urine<br>Volume | 2 (0.3%) | 2 | 0 | 0 | 0 | 2/2 |
| Hypercalciuria + Acid<br>Urine + Low Urine Volume | 6 (1.0%) | 5 | 0 | 1 | 0 | 5/6 |
| Hyperuricosuria + Low<br>Urine Volume | 4 (0.7%) | 4 | 0 | 0 | 0 | 3/4 |
| Hyperuricosuria + Acid<br>Urine | 7 (1.1%) | 5 | 2 | 0 | 0 | 5/7 |
| Hyperoxaluria +<br>Hyperuricosuria | 11<br>(1.8%) | 8 | 0 | 0 | 1 | 10/11 |
| Hypercalciuria +<br>Hyperuricosuria + Low<br>Urine Volume | 2 (0.3%) | 1 | 1 | 0 | 0 | 2/2 |
| Hypercalciuria +<br>Hyperuricosuria + Acid<br>Urine | 7 (1.1%) | 5 | 2 | 0 | 0 | 7/7 |
| Hyperuricosuria + Acid<br>Urine + Low Urine Volume | 2 (0.3%) | 2 | 0 | 0 | 0 | 2/2 |
| Hypercalciuria +<br>Hyperoxaluria +<br>Hyperuricosuria + Acid<br>Urine | 5 (0.8%) | 4 | 1 | 0 | 0 | 4/5 |
| Hyperoxaluria +<br>Hyperuricosuria + Low<br>Urine Volume | 1 (0.2%) | 1 (100%) | 0 | 0 | 0 | 1/1 |
| Hyperoxaluria +<br>Hyperuricosuria + Acid<br>Urine | 4 (0.7%) | 3 (75%) | 0 | 1 (25%) | 0 | 4/4 |
| Hypercalciuria +<br>Hyperoxaluria + Acid<br>Urine + Low Urine Volume | 4 (0.7%) | 3 (75%) | 0 | 1 (25%) | 0 | 4/4 |
| Hypercalciuria +<br>Hyperuricosuria + Acid<br>Urine + Low Urine Volume | 2 (0.3%) | 1 (50%) | 0 | 1 (50%) | 0 | 1/2 |
| Hypercalciuria +<br>Hyperoxaluria +<br>Hyperuricosuria + Low<br>Urine Volume | 1 (0.2%) | 1 (100%) | 0 | 0 | 0 | 1/1 |
| Hyperoxaluria +<br>Hyperuricosuria + Acid<br>Urine + Low Urine Volume | 1 (0.2%) | 1 (100%) | 0 | 0 | 0 | 1/1 |
| Hypercalciuria +<br>Hyperoxaluria +<br>Hyperuricosuria + Acid<br>Urine + Low Urine Volume | 1 (0.2%) | 1 (100%) | 0 | 0 | 0 | 1/1 |

## References

[1] Antonelli JA, Maalouf NM, Pearle MS, Lotan Y. Use of the National Health and Nutrition Examination Survey to Calculate the Impact of Obesity and Diabetes on Cost and Prevalence of Urolithiasis in 2030. European Urology 2014;66:724–9. doi:10.1016/j.eururo.2014.06.036.

[2] Geraghty RM, Cook P, Walker V, Somani BK. Evaluation of the economic burden of kidney stone disease in the UK: a retrospective cohort study with a mean follow-up of 19 years. BJU Int 2020;125:586–94. doi:10.1111/bju.14991.

[3] Lien C-S, Huang C-P, Chung C-J, Lin C-L, Chang C-H. Increased risk of anxiety among patients with urolithiasis: A nationwide population-based cohort study. Int J Urol 2015;22:937–42. doi:10.1111/iju.12865.

[4] Canales BK, Sharma N, Yuzhakov SV, Bozorgmehri S, Otto BJ, Bird VG. Long-term Recurrence Rates in Uric Acid Stone Formers With or Without Medical Management. Urology 2019;131:46–52. doi:10.1016/j.urology.2019.05.023.

[5] Ljunghall S, Danielson BG. A Prospective Study of Renal Stone Recurrences. British Journal of Urology 1984;56:122–4. doi:10.1111/j.1464-410X.1984.tb05346.x.

[6] Kazemi Y, Mirroshandel SA. A novel method for predicting kidney stone type using ensemble learning. Artif Intell Med 2018;84:117–26. doi:10.1016/j.artmed.2017.12.001.

[7] Moreira DM, Friedlander JI, Hartman C, Elsamra SE, Smith AD, Okeke Z. Using 24-hour urinalysis to predict stone type. J Urol 2013;190:2106–11. doi:10.1016/j.juro.2013.05.115.

[8] Boulesteix AL, Schmid M. Machine learning versus statistical modeling. Biometrical Journal 2014;56:588–93. doi:10.1002/bimj.201300226.

[9] Rule AD, Lieske JC, Li X, Melton LJ, Krambeck AE, Bergstralh EJ. The ROKS nomogram for predicting a second symptomatic stone episode. J Am Soc Nephrol 2014;25:2878–86. doi:10.1681/ASN.2013091011.

[10] Iremashvili V, Li S, Penniston KL, Best SL, Hedican SP, Nakada SY. External Validation of the Recurrence of Kidney Stone Nomogram in a Surgical Cohort. J Endourol 2019;33:475–9. doi:10.1089/end.2018.0893.

[11] Türk C, Petřík A, Sarica K, Seitz C, Skolarikos A, Straub M, et al. EAU Guidelines on Interventional Treatment for Urolithiasis. European Urology 2016;69:475–82. doi:10.1016/j.eururo.2015.07.041.

[12] Pearle MS, Goldfarb DS, Assimos DG, Curhan G, Denu-Ciocca CJ, Matlaga BR, et al. AUA Guidelines Medical Management of Kidney Stones: AUA Guideline. Juro 2014;192:316–24. doi:10.1016/j.juro.2014.05.006.

[13] Eisner BH, Goldfarb DS. A Nomogram for the Prediction of Kidney Stone Recurrence. Journal of the American Society of Nephrology 2014:1–3. doi:10.1681/ASN.2013.111180.

[14] Hsi RS, Sanford T, Goldfarb DS, Stoller ML. The Role of the 24-Hour Urine Collection in the Prevention of Kidney Stone Recurrence. J Urol 2017;197:1084–9. doi:10.1016/j.juro.2016.10.052.

[15] Fink HA, Wilt TJ, Eidman KE, Garimella PS, MacDonald R, Rutks IR, et al. Medical management to prevent recurrent nephrolithiasis in adults: a systematic review for an American College of Physicians Clinical Guideline. Ann Intern Med 2013;158:535–43. doi:10.7326/0003-4819-158-7-201304020-00005.

[16] Walker V, Stansbridge EM, Griffin DG. Demography and biochemistry of 2800 patients from a renal stones clinic. Annals of Clinical Biochemistry 2013;50:127–39. doi:10.1258/acb.2012.012122.

[17] Dhayat NA, Lüthi D, Schneider L, Mattmann C, Vogt B, Fuster DG. Distinct phenotype of kidney stone formers with renal phosphate leak. Nephrol Dial Transplant 2019;34:129–37. doi:10.1093/ndt/gfy170.

[18] Dhayat NA, Schaller A, Albano G, Poindexter J, Griffith C, Pasch A, et al. The Vacuolar H+-ATPase B1 Subunit Polymorphism p.E161K Associates with Impaired Urinary Acidification in Recurrent Stone Formers. J Am Soc Nephrol 2016;27:1544–54. doi:10.1681/ASN.2015040367.

[19] Therneau TM, Atkinson B, Ripley MB. The rpart package 2010.

[20] Segal MR. Machine Learning Benchmarks and Random Forest Regression 2004.

[21] Stekhoven DJ. missForest: Nonparametric missing value imputation using random forest. Astrophysics Source Code Library n.d.:ascl:1505.011.

[22] Kuhn M. caret: Classification and Regression Training. Astrophysics Source Code Library n.d.:ascl:1505.003.

[23] Daudon M, Lacour B, Jungers P. Influence of body size on urinary stone composition in men and women. Urological Research 2006;34:193–9. doi:10.1007/s00240-006-0042-8.

[24] Yasui T, Iguchi M, Suzuki S, Kohri K. Prevalence and Epidemiological Characteristics of Urolithiasis in Japan: National Trends Between 1965 and 2005. Urology 2008;71:209–13. doi:10.1016/j.urology.2007.09.034.

[25] Eisner BH, Sheth S, Dretler SP, Herrick B, Pais VM. Abnormalities of 24-hour urine composition in first-time and recurrent stone-formers. Urology 2012;80:776–9. doi:10.1016/j.urology.2012.06.034.

[26] Curhan GC, Taylor EN. 24-h uric acid excretion and the risk of kidney stones. Kidney International 2008;73:489–96. doi:10.1038/sj.ki.5002708.

[27] Flannigan R, Choy WH, Chew B, Lange D. Renal struvite stones--pathogenesis, microbiology, and management strategies. Nat Rev Urol 2014;11:333–41. doi:10.1038/nrurol.2014.99.

[28] Chillarón J, Font-Llitjós M, Fort J, Zorzano A, Goldfarb DS, Nunes V, et al. Pathophysiology and treatment of cystinuria. Nat Rev Nephrol 2010;6:424–34. doi:10.1038/nrneph.2010.69.

[29] Geraghty R, Abdi A, Somani B, Cook P, Roderick P. Does chronic hyperglycaemia increase the risk of kidney stone disease? results from a systematic review and meta-analysis. BMJ Open 2020;10:e032094. doi:10.1136/bmjopen-2019-032094.

[30] Grases F, Costa-Bauzá A, Ramis M, Montesinos V, Conte A. Simple classification of renal calculi closely related to their micromorphology and etiology. Clin Chim Acta 2002;322:29–36. doi:10.1016/s0009-8981(02)00063-3.

[31] Gentle DL, Stoller ML, Bruce JE, Leslie SW. Geriatric urolithiasis. Juro 1997;158:2221–4. doi:10.1016/s0022-5347(01)68203-x.

[32] Fuster DG, Morard GA, Schneider L, Mattmann C, Lüthi D, Vogt B, et al. Association of urinary sex steroid hormones with urinary calcium, oxalate and citrate excretion in kidney stone formers. Nephrol Dial Transplant 2020. doi:10.1093/ndt/gfaa360.

[33] Yamout H, Goldberg S. Genetic and Environmental Risk Factors for Kidney Stones. Nutritional and Medical Management of Kidney Stones, vol. 62, Cham: Humana, Cham; 2019, pp. 43–52. doi:10.1007/978-3-030-15534-6_3.

[34] Hiatt RA, Ettinger B, Caan B, Quesenberry CP, Duncan D, Citron JT. Randomized controlled trial of a low animal protein, high fiber diet in the prevention of recurrent calcium oxalate kidney stones. Am J Epidemiol 1996;144:25–33.

[35] Fink HA, Akornor JW, Garimella PS, MacDonald R, Cutting A, Rutks IR, et al. Diet, fluid, or supplements for secondary prevention of nephrolithiasis: a systematic review and meta-analysis of randomized trials. European Urology 2009;56:72–80. doi:10.1016/j.eururo.2009.03.031.

[36] Halbritter J, Baum M, Hynes AM, Rice SJ, Thwaites DT, Gucev ZS, et al. Fourteen monogenic genes account for 15% of nephrolithiasis/nephrocalcinosis. J Am Soc Nephrol 2015;26:543–51. doi:10.1681/ASN.2014040388.

[37] Paranjpe I, Tsao N, Judy R, Paranjpe M, Chaudhary K, Klein D, et al. Derivation and Validation of Genome Wide Polygenic Score for Urinary Tract Stone Diagnosis. Kidney International 2020. doi:10.1016/j.kint.2020.04.055.

[38] Howles SA, Wiberg A, Goldsworthy M, Bayliss AL, Gluck AK, Ng M, et al. Genetic variants of calcium and vitamin D metabolism in kidney stone disease. Nat Commun 2019;10:5175–10. doi:10.1038/s41467-019-13145-x.

[39] Eyre KS, Lewis F, Cui H, Grout E, Mihai R, Turney BW, et al. Utility of blood tests in screening for metabolic disorders in kidney stone disease. BJU Int 2020. doi:10.1111/bju.15250.

[40] Hill F, Sayer JA. Precision medicine in renal stone-formers. Urolithiasis 2019;47:99–105. doi:10.1007/s00240-018-1091-5.

[41] Sayer JA, Moochhala SH, Thomas DJ. The Medical Management of Urolithiasis:. British Journal of Medical and Surgical Urology 2010;3:87–95. doi:10.1016/j.bjmsu.2010.02.004.

